# A Systematic Review on Malaria and Tuberculosis (TB) Vaccine Challenges in Sub-Saharan African Clinical Trials

**DOI:** 10.1101/2024.02.28.24302787

**Authors:** Maonezi Abas Hamisi, Nur Ain Mohd Asri, Aini Syahida Mat Yassim, Rapeah Suppian

**Author notes:** Corresponding author: Maonezi Abas Hamisi, School of Science and Technical Education (CoSTE), Mbeya University of Science and Technology, Tanzania, P.O. Box 131 Mbeya, Tanzania.

## Abstract

**Objective:** For more than a century, developing novel and effective vaccines against malaria and tuberculosis (TB) infections has been a challenge. This review sought to investigate the reasons for the slow progress of malaria and TB vaccine candidates in sub-Saharan African clinical trials.

**Methods:** The systematic review protocol was registered on PROSPERO on July 26, 2023 (CRD42023445166). The research articles related to the immunogenicity, efficacy, or safety of malaria or TB vaccines that were published between January 1, 2012, and August 31, 2023, were searched on three databases: Web of Science (WoS), PubMed, and ClinicalTrials.gov.

**Results:** A total of 2342 articles were obtained, 50 of which met the inclusion criteria. 28 (56%) articles reported on malaria vaccine attributes, while 22 (44%) articles reported on TB vaccines. In both cases, the major challenges in sub-Saharan African clinical trials were immunogenicity and efficacy, rather than safety.

**Conclusion:** Factors such as population characteristics, pathogen genetic diversity, vaccine nature, strategy, and formulation were associated with slow progress of the malaria and TB vaccine candidates in sub-Saharan African clinical trials.

**Author summary:** Vaccines are one of the most powerful control strategies for both infectious and non-infectious diseases. The lack of durable vaccines and the development of antimicrobial resistance have made malaria and TB threats to human lives, specifically in sub-Saharan Africa. The search for novel, reliable, and durable vaccines against these infections has been underway for more than a decade. Despite this, none have resulted in vaccines with all three preferred critical attributes: immunogenicity, efficacy, and safety. This indicates that there are hindering challenges that have been neglected in the development pipeline. This review focused on understanding these challenges in sub-Saharan African clinical trials. The results of the study are crucial for the identification of areas for improvement in vaccine design, evaluation, and implementation strategies, hence driving advancements in NTD vaccine research and development.

## 1. Introduction

Malaria and Tuberculosis (TB) are among the top ten causes of death in low- and middle-income countries, the majority of which are sub-Saharan countries (1). Malaria is a plasmodium-borne infection that is spread by female Anopheles mosquitoes (2). *Plasmodium falciparum* species are the dominant cause of human malaria (3). In 2021, 619,000 deaths were caused by malaria worldwide, 95% of which occurred in sub-Saharan Africa (4). The favourable environment for *P. falciparum* species (5) and healthcare systems contribute to this prevalence (6). In contrast, human TB is an airborne disease caused by *Mycobacterium tuberculosis* (Mtb). TB spreads through respiratory system encounters active Mtb-containing air droplets (7). TB infections were the leading cause of human deaths prior to COVID-19 (7). Human TB causes 10.6 million cases and 1.6 deaths, 90% of which occurred in sub-Saharan Africa (7).

Currently, chemical drugs, as well as vaccines, are used for the control of malaria (8) and TB (7). However, the development of resistant strains has made drug use inefficient and costly. Vaccines are the most effective options for these diseases, and they can help to prevent the spread of resistance. Despite not being fully certified, the malaria RTS, S/AS01, was recommended for the pilot vaccination of 5- to 17-month-old children living in high-endemic areas(9). Unlike malaria, BCG is the only certified TB vaccine. Both RTS, S/AS01 (10), and BCG (11) share similar limitations: they provide protection but limited to young age groups. Different groups of malaria vaccine candidates have been tested in sub-Saharan African clinical trials. Some of these include subunit vaccines (12,13), (14), (15,16), viral-like particle vaccines (17–20), and whole attenuated vaccines (21–23). Like malaria vaccine candidates, TB vaccine candidates include subunit vaccines (24–27), (28–30), (31–34), inactivated vaccines (35), and whole attenuated vaccines (11,36,37). Nevertheless, the progress of malaria and TB vaccine development in sub-Saharan Africa has been arduous and frequently regarded as sluggish. The aim of this study was to elucidate the barriers that hinder the rapid advancement of efficacious malaria and TB vaccines, with a specific emphasis on immunogenicity, effectiveness, and safety in sub-Saharan Africa.

## 2. Methodology

The review protocol was registered on PROSPERO (CRD42023445166). In brief, this review followed the Preferred Reporting Items for Systematic Reviews and Meta-Analysis (PRISMA) guidelines (38). The keywords and review questions were formulated based on PICO: Sub-Saharan Africans (population), malaria and TB vaccine candidates (intervention), non-vaccinated, placebo, or any control setting (control), immunogenicity, efficacy, and safety (outcomes). The review question was “What are the challenges that hinder the rapid development of malaria and TB vaccines in sub-Saharan African clinical trials?”.

### 2.1 Article identification

The review began with the identification of keywords and their respective synonyms by the first and second authors (HM and NA, respectively). The keywords and synonyms used to formulate the search strategy were based on the PICO formulation, which includes Sub-Saharan Africans (population), malaria and TB vaccine candidates (intervention), non-vaccinated, placebo, or any control setting (control), immunogenicity, efficacy, and safety (outcomes) as summarized in **Table 1**. The search was performed between the 1st of August 2023 and the 31st of August 2023 on three databases, Web of Science, PubMed, and ClinicalTrials.gov, with some refinement to meet the inclusion criteria.

**Table 1.**
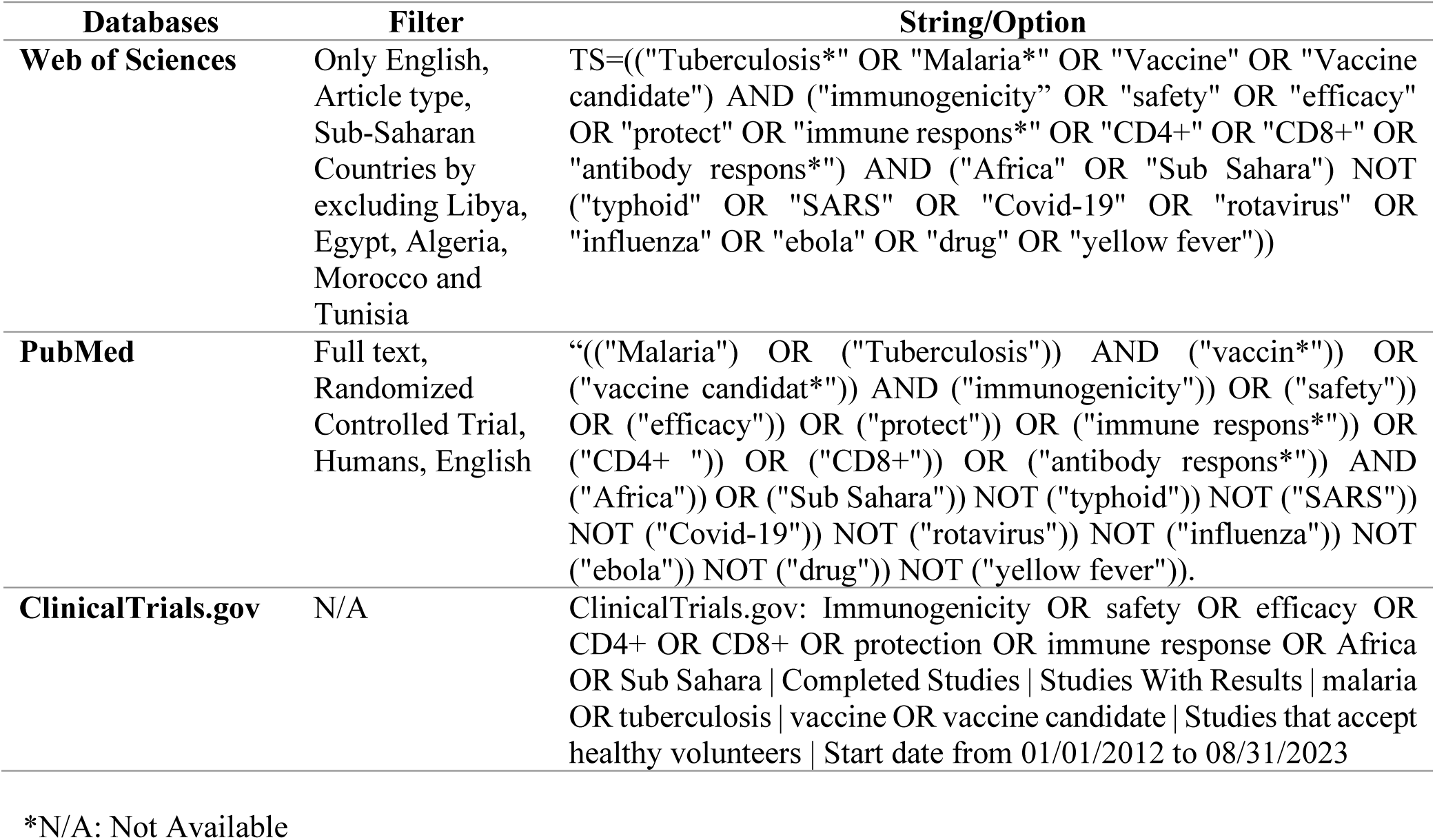
The search strategy applied in this study.

| Databases | Filter | String/Option |
| --- | --- | --- |
| **Web of Sciences** | Only English,<br>Article type,<br>Sub-Saharan<br>Countries by<br>excluding Libya,<br>Egypt, Algeria,<br>Morocco and<br>Tunisia | TS=((("Tuberculosis*" OR "Malaria*" OR "Vaccine" OR "Vaccine candidate") AND ("immunogenicity" OR "safety" OR "efficacy" OR "protect" OR "immune respons*" OR "CD4+" OR "CD8+" OR "antibody respons*") AND ("Africa" OR "Sub Sahara") NOT ("typhoid" OR "SARS" OR "Covid-19" OR "rotavirus" OR "influenza" OR "ebola" OR "drug" OR "yellow fever")) |
| **PubMed** | Full text,<br>Randomized<br>Controlled Trial,<br>Humans, English | ((("Malaria") OR ("Tuberculosis")) AND ("vaccin*")) OR ("vaccine candidat*") AND ("immunogenicity")) OR ("safety")) OR ("efficacy")) OR ("protect")) OR ("immune respons*")) OR ("CD4+ ") OR ("CD8+")) OR ("antibody respons*")) AND ("Africa")) OR ("Sub Sahara")) NOT ("typhoid")) NOT ("SARS")) NOT ("Covid-19")) NOT ("rotavirus")) NOT ("influenza")) NOT ("ebola")) NOT ("drug")) NOT ("yellow fever"))). |
| **ClinicalTrials.gov** | N/A | ClinicalTrials.gov: Immunogenicity OR safety OR efficacy OR CD4+ OR CD8+ OR protection OR immune response OR Africa OR Sub Sahara \| Completed Studies \| Studies With Results \| malaria OR tuberculosis \| vaccine OR vaccine candidate \| Studies that accept healthy volunteers \| Start date from 01/01/2012 to 08/31/2023 |

\*N/A: Not Available

### 2.2 Article screening and eligibility

Two authors, HM, and NA screened the articles obtained independently. Malaria or TB vaccine candidate studies that reported immunogenicity, efficacy, safety, or a combination, involved a sub-Saharan population, were randomised clinical trials, published in English between January 1, 2012, and August 31, 2023, were included. Studies not meeting these criteria were excluded. The discrepancies occurred were resolved by involving the third and fourth authors (AS and RS). The duplicate and irrelevant articles were removed using Mendeley software (2.107.0, 2023).

### 2.3 Assessment of study quality

The risk of bias in the eligible studies was analysed by the first author (HM) using the Cochrane risk of bias tool (RoB 2)(38). The other three co-authors confirmed the findings (NA, AS, and RS). Each randomised controlled trial was rated as “high,” “low,” or “some concerns” for bias in five domains: randomization process, deviation from intervention, missing outcome data, measurement of outcomes, and selection of reported outcomes.

### 2.4 Data extraction and synthesis

Data were extracted and compiled from all eligible articles on the standard data collection table. The first author’s surname, publication year, population, region (country), clinical trial phase, intervention, control, and number of doses were retrieved for the articles. We then performed a narrative synthesis of the literature using our outcome keywords immunogenicity, efficacy, and safety.

## 3. Results

### 3.1 Study selection

The search yielded 2342 publications. 1791 (76.5%) articles were from Web of Sciences, 497 (21.2%) from PubMed, and 54 (2.3%) from ClincalTrials.gov. The ClincalTrials.gov database contained 54 articles from 11 studies (55%) of 20 search projects. To minimise bias, nine (45%) studies had no publications and were excluded from this review. The 2342 published studies were reduced to 92 (3.9%) duplicates and 2179 (93%) irrelevant. 17 (23.9%) studies were removed after screening titles and abstracts of 71 (3.0%). After full-text screening, 4 (7.4%) of 54 (76.1%) studies were removed. Fifty studies were eligible for review; 28 (56%) were malaria vaccines and 22 (44%) were TB vaccines. The flowchart of this study was summarized in **Figure 1** and **Table 2** highlighted all eligible studies’ characteristics.

**Figure 1.**
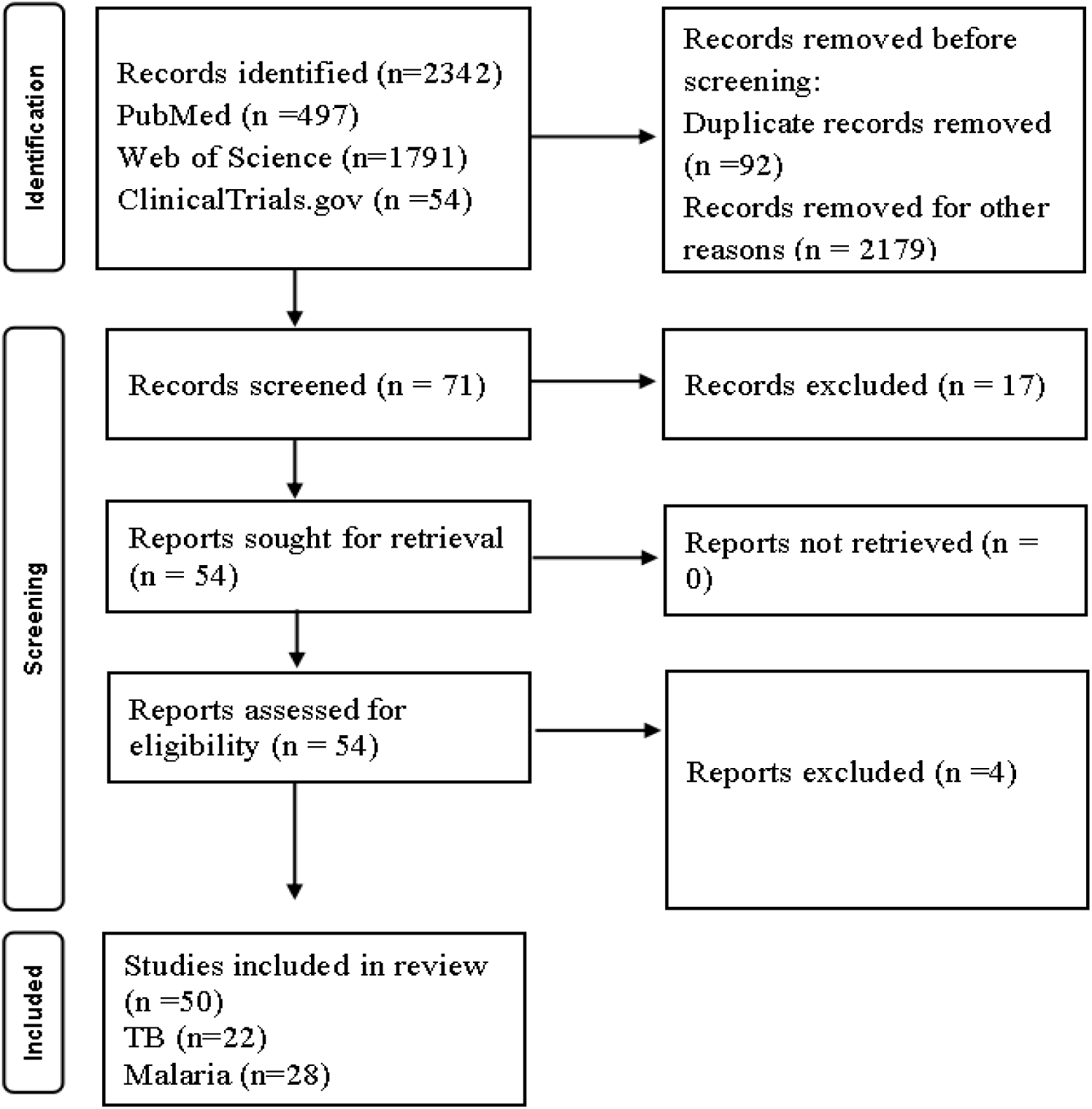
PRISMA flowchart of this study.

**Table 2.**
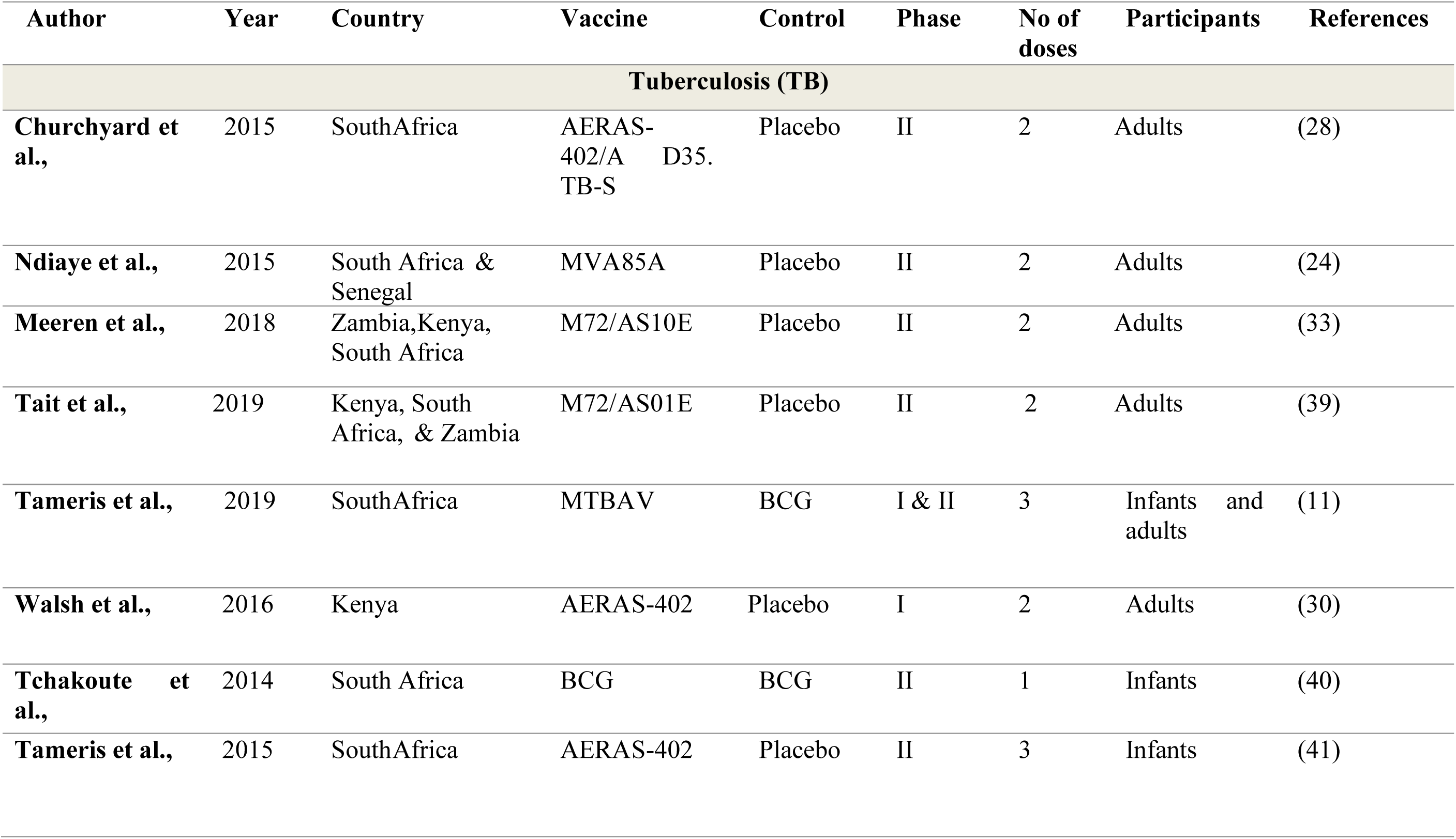
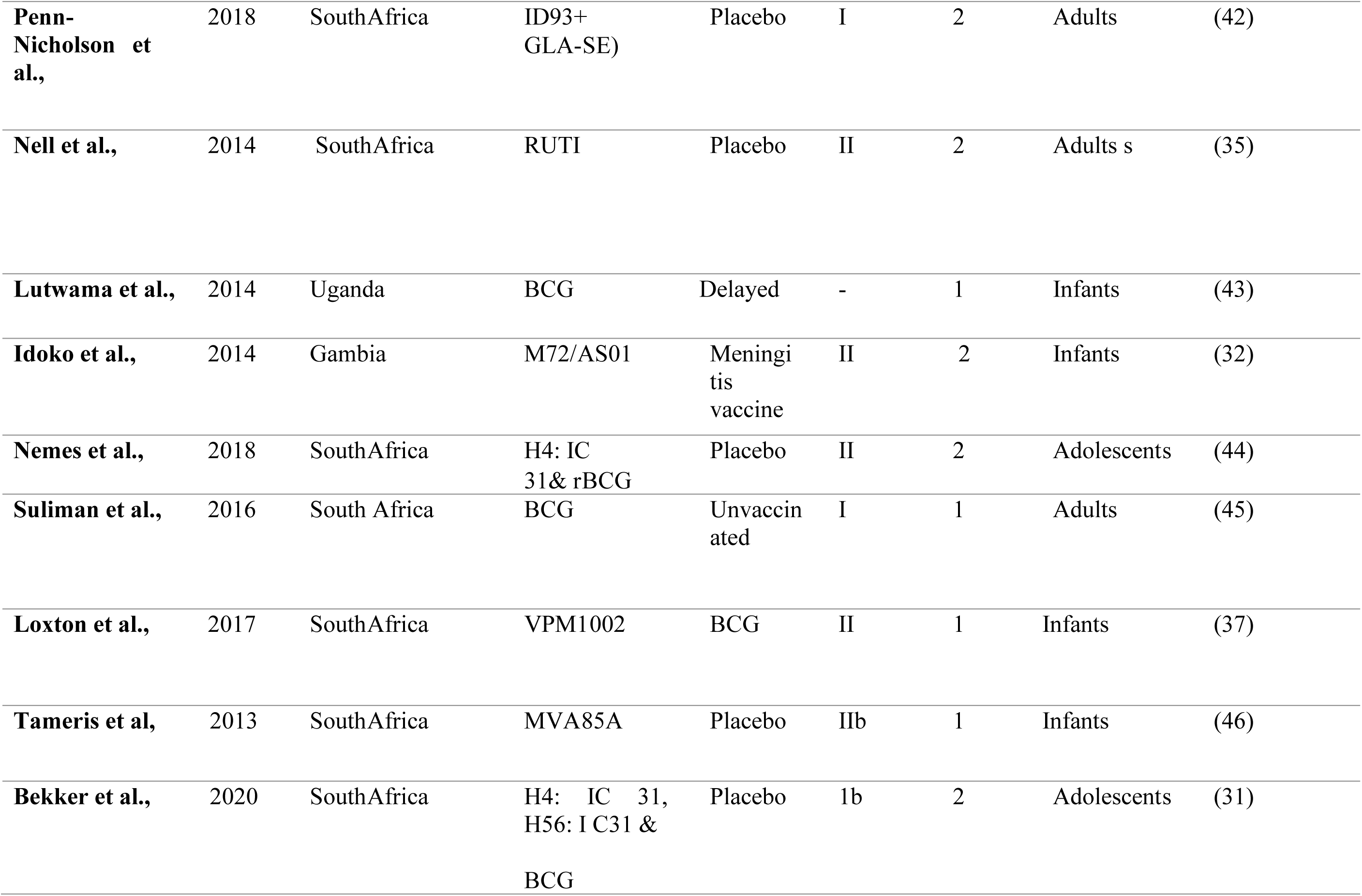
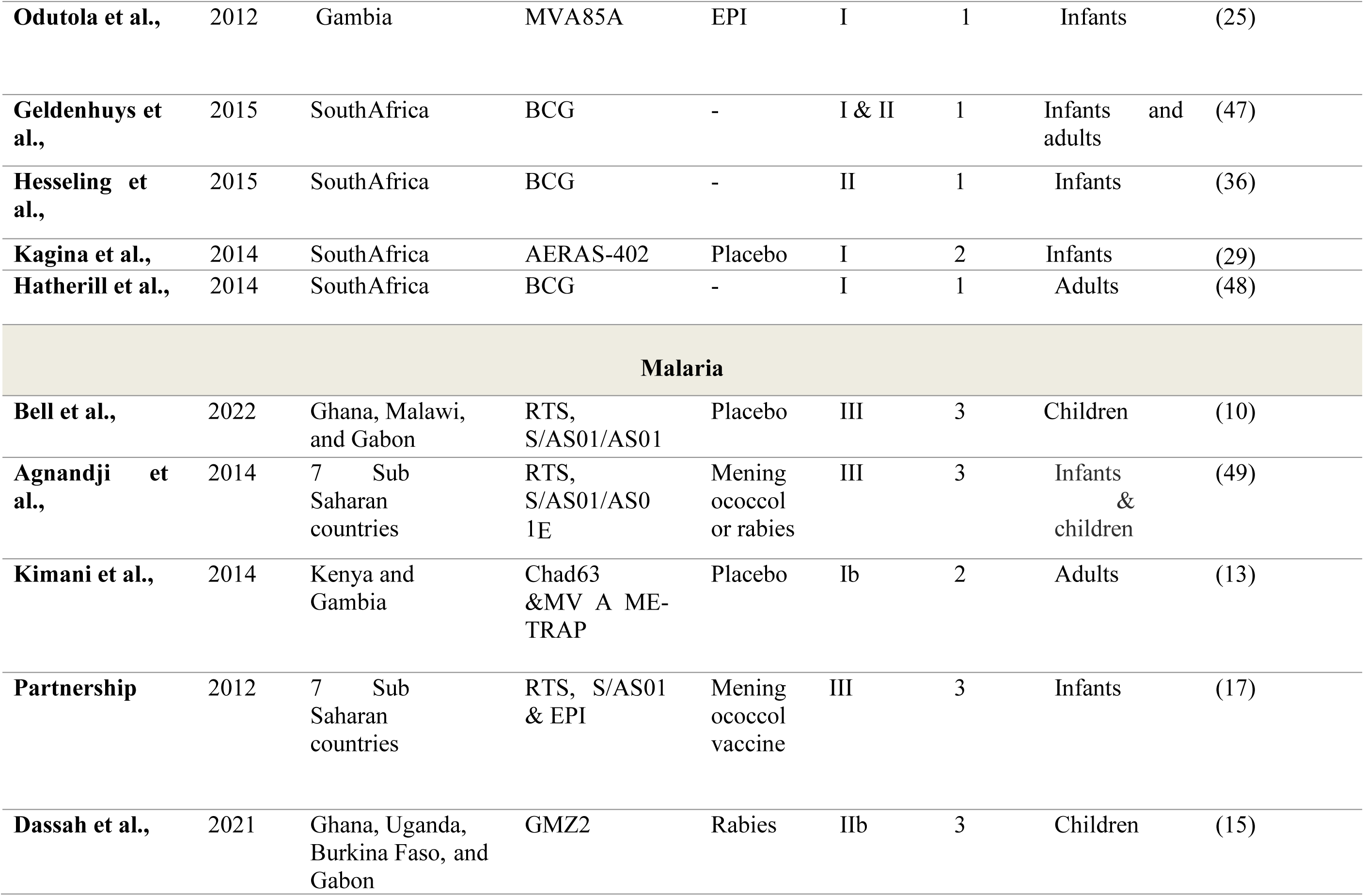
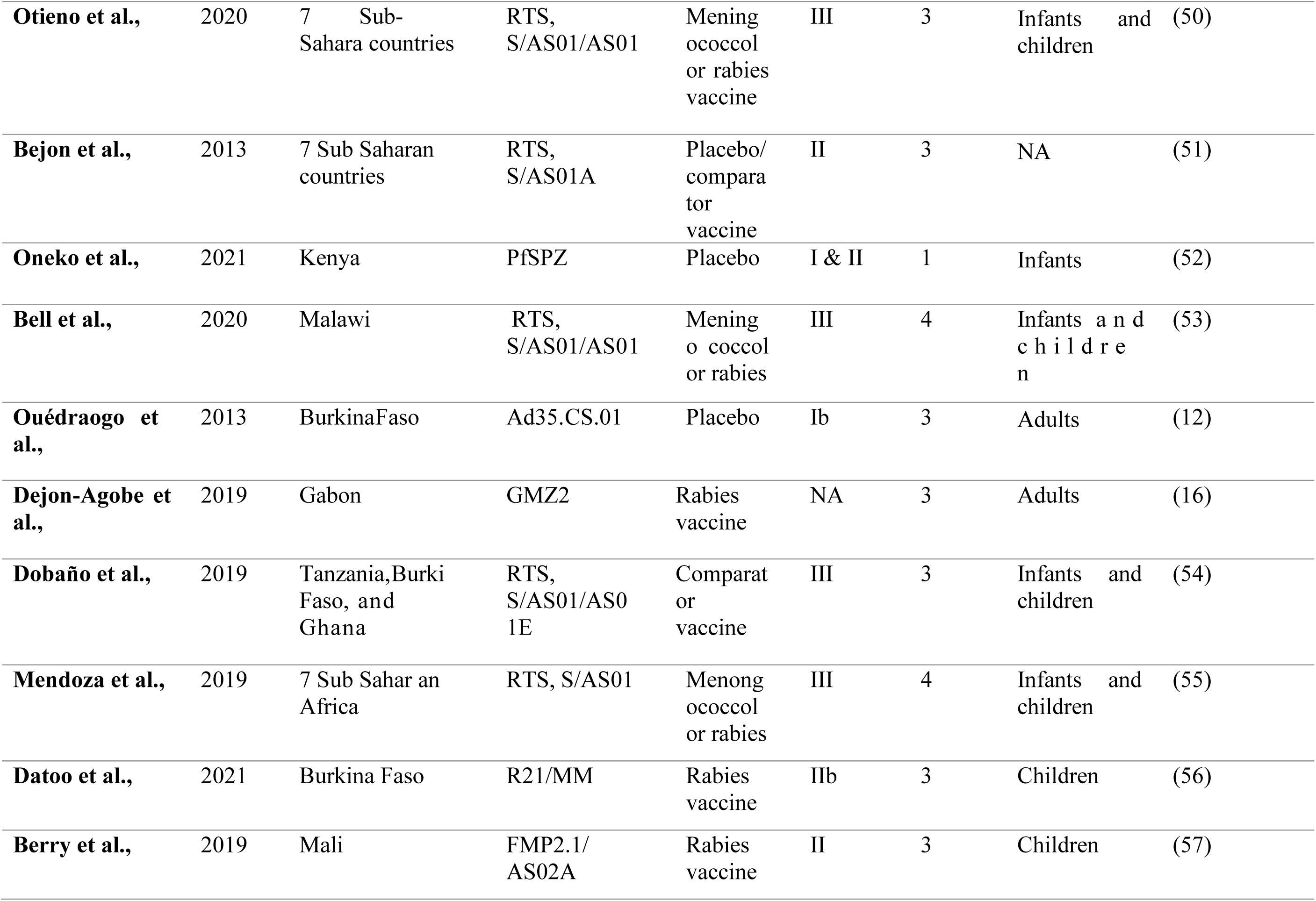
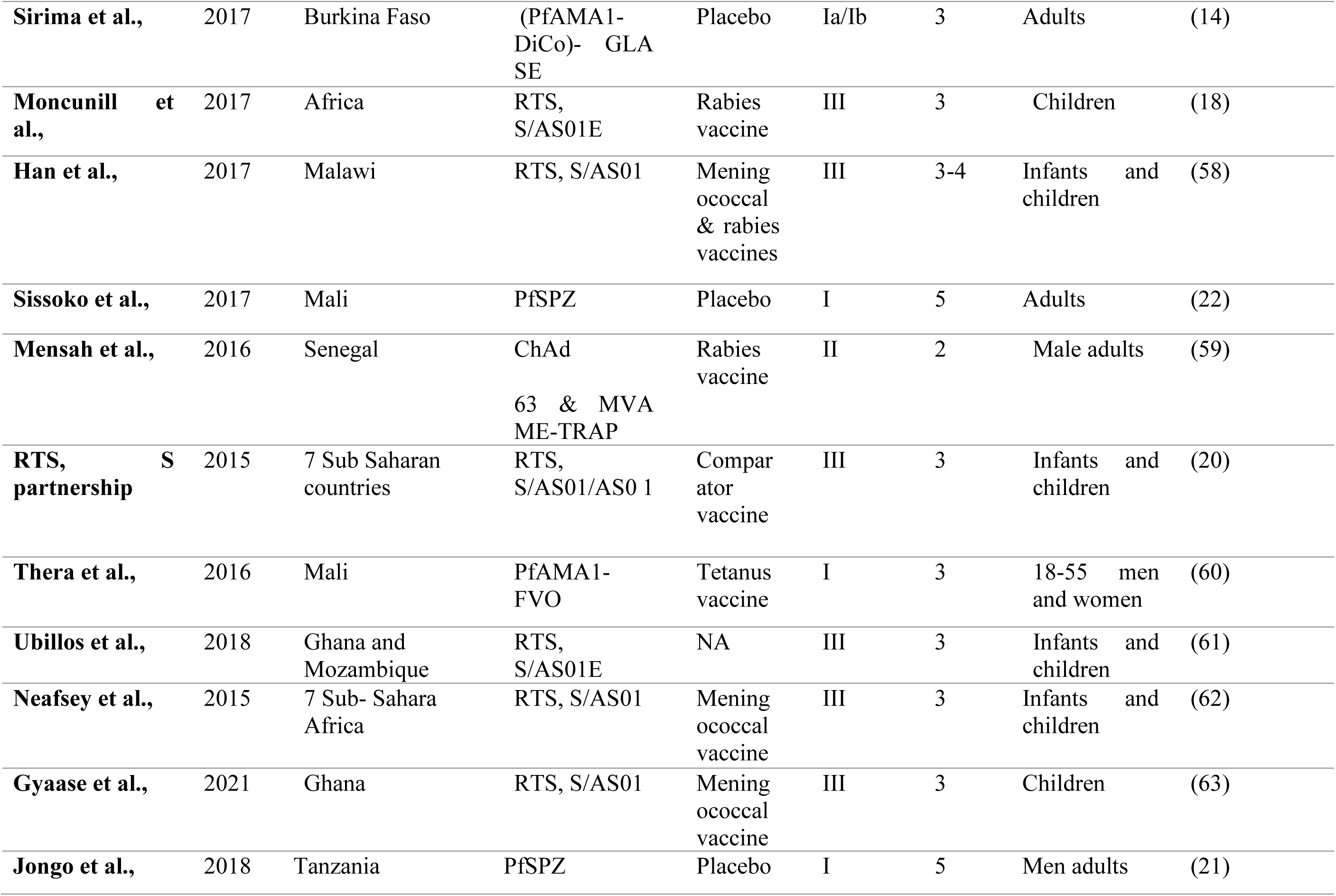
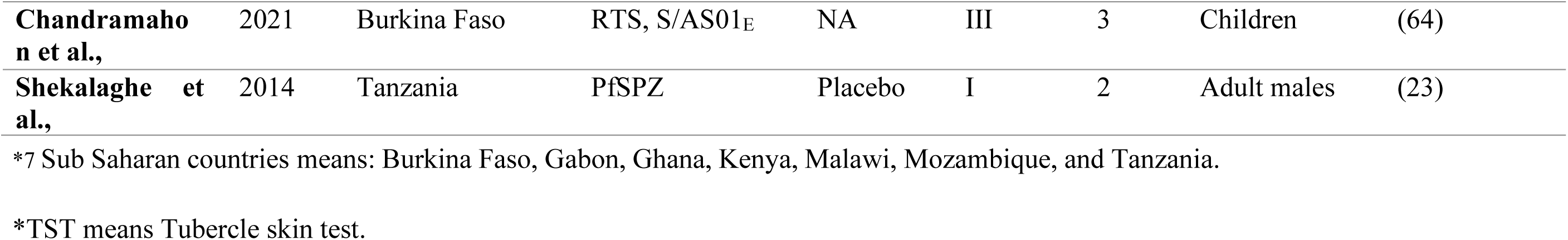
Characteristics of all eligible studies in this review.

### 3.2 Quality assessment

The overall quality of all 50 eligible studies was good. 47 (94%) studies received “low risk” overall status for causing bias, 3 (6%) studies received “some concerns” status (13,18,48), and there were no articles with “high risk” overall status (**Figure 2**).

**Figure 2.**
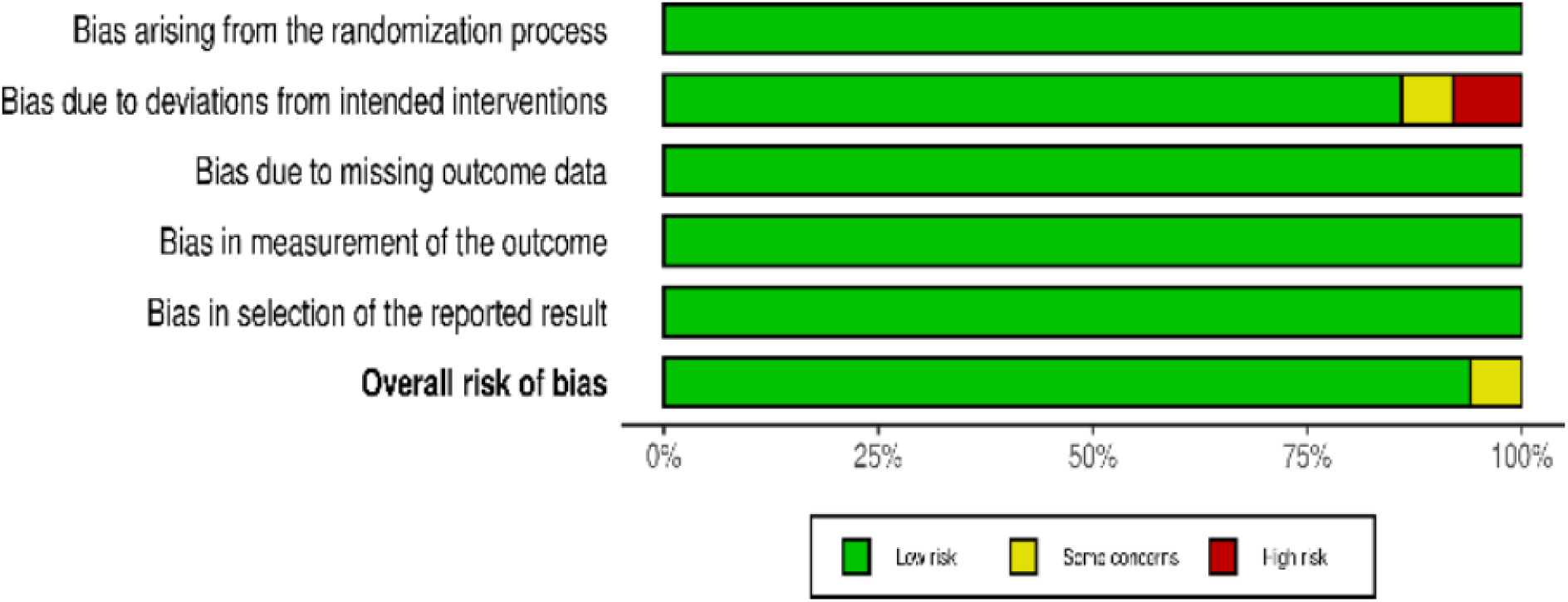
Robvis risk assessment summary for all randomised controlled trials in this review.

### 3.3 Immunogenicity

Immunogenicity varied in magnitude, duration, and type for most malaria and TB vaccine candidates. Most malaria vaccine candidates had seven times the immunogenicity of humoral immune cells compared to CD4+ and CD8+ cells, which appeared two times (7.14%). Conversely, TB vaccine candidates were more immunogenic to CD4+ T cells (72.7%) than CD8+ T cells (45.50%). Most TB vaccine candidates induce CD4+ and CD8+ cells only (24,29,35,37,43,45,47), followed by CD4+, CD8+ cells and antibodies only (28,30,31,41) and CD4+ cells and antibodies only (32,42). Additionally, very few candidates induce only CD4+ cells (11,25,40), and there are no vaccine candidates that induce CD8+ cells only. These findings demonstrate the immunogenic diversity of malaria and TB vaccine candidates, emphasising the need for tailored vaccine design and evaluation.

### 3.4 Efficacy

This review revealed that out of 28 malaria-related studies, only the R21/MM vaccine candidate (3.8%) had the highest vaccine efficacy (VE) of 77% (56), followed by RTS, S/AS0, 50.3% VE (62), 46% VE (49), 45% VE (51), 28% VE (20), 27% VE (49), 12% VE (52), and 18% VE (20), as well as 8% VE (59) in adults. Other malaria-related studies reported vaccine efficacy qualitatively (22,63,64). On the other hand, the M72/AS01E TB vaccine candidate showed the highest vaccine efficacy, 54.0% VE (33), followed by 49.7% VE (39), 45.4% VE, and 30.5% VE (44). In some cases, MVA85A vaccine candidates have shown insignificant efficacy (24,27). Certain candidates show promise as disease-fighting tools due to their efficacy. However, insignificant efficacy in some cases highlights the need for continued research and development to improve malaria and TB vaccines.

### 3.5 Safety

This review revealed that both malaria and TB vaccine candidates had no safety issues that resulted in the termination of the studies. The common localized adverse effects (AEs) of the malaria vaccine candidates included injection site swelling (9, 32.14%) and pain (8, 28.6%), whereas the systemic AEs included headache (12, 42.9%), myalgia, fever, and nausea (8, 28.6%). The severe adverse effects (SAEs) included malaria (8, 28.6%), meningitis, and febrile convulsion (7; 25%). In contrast, the common AEs associated with TB vaccine candidates were injection site pain (5, 22.7%) and systemic abdominal pain (11, 50.0%). The SAEs were gastroenteritis (3, 13.6%). This study found that vaccine candidates for malaria and TB may cause some side effects, but the risks are generally tolerable. No studies were stopped due to safety concerns.

## 4. Discussion

The systematic review of 50 studies highlights immunogenicity and efficacy as major challenges in sub-Saharan African malaria and TB vaccine clinical trials. Safety was not a challenge. However, this factor should not be ignored in new vaccine development against these diseases since factors such as dose (23,34,52), vaccine type (44), age group (55), and health status (28) have some impact on safety. It is critical to evaluate the factors that contribute to variations in immunogenicity and efficacy during the development of vaccines against malaria and TB, especially in clinical trial settings. Complexities arise because of factors such as population attributes, vaccine strategy, and pathogen genetic diversity, all of which influence the immune responses of vaccine candidates. Thus, given the complexity of the factors, it appears that no general conclusion can be drawn regarding the variation.

Lack of generalizability, which considered as limitation of evidence in this review, necessitates that each factor be examined individually. We discovered that population characteristics play the most important role in variation of immunogenicity and efficacy during malaria and TB vaccine clinical trials. There is a correlation between age group and vaccine candidate protection. For example, non-cytophilic antibodies increase the risk of malaria infection while also reducing the immunogenicity and efficacy of RTS and S/AS01E (61). These antibodies may also explain why malaria is more common in older people (65). However, the complexity arises when the effects of these antibodies were found to be more immunogenic in malaria-naïve adults than in semi-immune adults (13). This finding suggests that infection exposure also contributes to the immunogenicity and efficacy variations. This was supported by findings that high malaria transmission intensity reduced the efficacy of RTS, S/AS01 efficacy (10,51). Ideally, older age groups might have more non-cytophilic antibodies than younger age groups due to the longer exposure time. Immunogenicity and efficacy improved more in older vaccinees than young ones (15,20,49,58). Furthermore, factors such as lower socioeconomic status (SES) (63), forest ecological context (53), and season (64) in the respective region were associated with low efficacy. This contributes to the variations in immunogenicity and efficacy that are caused by population characteristics.

The vaccine strategy significantly influences the immunogenicity and effectiveness of potential vaccines. This review observes the impact of BCG vaccination on infants at various time points and found mixed results of TB protection. Vaccination at birth results in more distinct cellular immune responses than vaccination at 6 weeks (43) or vaccination at 14 weeks (36). In contrast, there was no significant difference in the prevalence of BCG vaccination at birth or at 8 weeks after delivery. However, there is a subsequent shift in this response, as infants who were delayed in their attempts to receive the vaccine demonstrate increased cellular immune responses. These changes could potentially be attributed to HIV exposure at birth, a period when the immune system lacks the capability to combat the viruses (40). Thus, vaccination timepoints affect vaccine candidate immune response.

This review revealed that the inferior performance of the vaccine candidates was associated with either poor immunogenicity or a narrow range of immune response inductions. For example, a vaccine study in which H56:IC31, H4:IC31, and BCG were used induced high antigen specific CD4+ and IgG expression. However, the number of CD8+ cells induced was very low (31). Boosting with BCG enhanced only anti-BCG CD4+ but not anti-H4 or anti-H56 IgG antibodies. Similarly, the BCG booster of MVA85A failed to enhance anti-Ag85A immune responses (44). The induction of a few immune responses affects the treatment efficacy. Anti-CSP and anti-HB-specific IgG as well as IgM antibodies defined the efficacy of the RTS and S/AS01 vaccine candidates (61). Despite being different from one endemic region to another, the efficacy of these agents decreased once the anti-HB antibodies were not induced (17). Additionally, the lack of PfSPZ vaccine candidate efficacy at 6 months postimmunization was associated with the lack of induction of cellular T immune responses (52). This showed that multiple immune responses must work together for robust protection. This factor relates to the vaccine strategy to improve malaria and TB vaccine candidates’ immunogenicity and efficacy in clinical trials.

Pathogen genetic diversity affects vaccine immunogenicity and efficacy in clinical trials. Allelic mismatches between vaccine antigens and natural plasmodium species affect immunogenicity and efficacy (62). In addition, there were more than 90% mismatches in Sub-Saharan Africa. This might be another major reason for the diminishing protection efficacy (12,50,58). However, this review revealed that boosting with additional doses (53,56) and the use of a strong adjuvant (51) might prevent waning protection. However, continuous boosting might not be useful for low- or middle-income countries (LMICs), such as Sub-Saharan Africa. Furthermore, adjuvant effects, such as RTS, S/AS01 and RTS, S/AS02A (51), GMZ2/alum (15), and GMZ2/Alhydrogel/liposomes (16), and AMA1-DiCo GLA-SE and AMA1-DiCo Alhydrogel (14), have been reported in comparative studies.

However, these similar differences were not observed during the GMZ2 Alhydorgel and GMZ2 CFA01 studies (16). Despite this, pre-evaluation of adjuvants for conjugation is crucial. Other vaccine candidates lacked correlation protection. For example, the FMP2.1/AS02A (57), GMZ2 (15,16), PfSPZ (21), and MVA-vectored TB candidates were immunogenic but not protective (24,27). This observation was like that of the Nouatin *et al*. study in Gabon (66). The lack of specificity may have played a role in these observations.

Most of the studies included in this review induced all immune responses (CD4+, CD8+, IgG, IgA, and IgM). Most CSP-based malaria vaccine candidates induce a greater quantity of anti-NANP antibodies than anti-C-terminal antibodies (54,56). This might be because the NANP region is conserved, while the C-terminus is polymorphic (67). The authors added that removing the antigen parts responsible for the induction of non-neutralizing antibodies might reduce this challenge. However, the profile of antibodies and CD8+ cells induced by Ad.35.CS.01 was based on the CSP and Ad35 antigens (12). In HIV-infected vaccinees, the reduction in immune responses was dose dependent (24,28,29). In some cases, the dose had no effect on immunogenicity (42). Other vaccine candidates, such as AERAS-402 in infants (41) and MTBVAC in adults and neonates (11), induce low immune responses at all vaccine dose levels. However, the highest MTBVAC dose was superior to the equivalent BCG dose. The three H56:IC31 doses were found to be highly immunogenic (68). In contrast, the highest dose of the RUTI vaccine candidate lost its immunogenicity capability (35). Furthermore, PfSPZ vaccination in infants induces significant humoral and insignificant cellular responses (52).

This review revealed that M72/AS01E was the most protective TB vaccine candidate (33). However, another study reported that M72/AS01E had decreased vaccine efficacy with no worsening tendency (39). These differences were associated with vaccine nature, vaccination strategies, genetic differences, and health status. The differences in nature of the vaccines used were as follows: BCG and H4:IC31 (44,69) and H56:IC31, H4:IC31, and BCG (31). Additionally, the immune responses induced during comparative RTS and S/AS01 vaccination between HIV and non-HIV vaccinees were greater in healthy individuals than in HIV vaccinees (50). These findings are consistent with a previous study in which HIV vaccinees were shown to reduce immune responses (19). The reduced immune responses in HIV-infected participants might be due to the ability of HIV to cause immune response dysfunction. Concomitant vaccination with antiretroviral drugs (ARVs) might reduce the impacts of these viruses [19].

Unfortunately, this review revealed that none of the studies employed mucosal delivery routes. The mucosal route enhances the speed and efficiency of both localized and systemic humoral and cellular immune responses, as well as innate and adaptive immune responses. The mucosal MVA85A immunization in the United Kingdom induced strong immunogenicity, which has not been observed in South Africa through traditional routes (70). Even though the natural route to malaria infection does not involve mucosal surfaces, mucosal immunization with malaria-inducing antibodies protects mice (71). In addition to the immunological point of view, mucosal routes may be more economical than parenteral routes because they do not require trained personnel and hence are appropriate in resource-limited sub-Saharan Africa.

The review acknowledges the absence of comparison groups and inconsistencies in observations across studies as limitations of the review processes. Furthermore, comprehensive interpretation of the findings is complicated by divergences in vaccine administration timing, dosage, and the existence of confounding variables such as HIV exposure. The ineffectiveness of specific vaccine candidates can be attributed to their inadequate immunogenicity, which means they elicit a restricted spectrum of immune responses. This underscores the criticality of refining the process of vaccine development and assessment. The review emphasises the importance of comprehensive immune responses, which comprise humoral and cellular components, to provide strong protection against malaria and TB. Further investigation is warranted to tackle the identified obstacles, including but not limited to optimising dosing regimens, enhancing the immunogenicity of vaccines that is specific to certain endemic region, and investigating alternative delivery routes such as mucosal administration to enhance vaccine efficacy, particularly in settings with limited resources like sub-Saharan Africa.

## 5. Conclusion

This review revealed that immunogenicity and efficacy are the major challenges for both malaria and TB vaccine candidates. The challenges were orchestrated by population characteristics, vaccination strategies, and pathogen genetic diversity. This review suggested that the continued neglect of these factors could lengthen the journey toward robust vaccines, that mucosal routes may improve vaccine candidate performance, and that the development of endemic region-based vaccines is worthwhile.

## Data Availability

All data produced in the present work are contained in the manuscript

## 8. Ethical approval and consent to participate

Note applicable.

## 9. Availability of data and material

All data and materials are available in this manuscript.

## 10. Author contribution

MH conceptualised and designed the study, collected, and interpreted data, and drafted the work. NA contributed to study conception, design, collection, interpretation, and work drafting. AS contributed to the data collection, interpretation, and drafting of the work. Furthermore, RS contributed to the conceptualization of the study, data collection, and interpretation. All authors revised and approved the final version. Also, the authors agreed to be accountable for the contribution made and the resolution of any questions that would arise about data accuracy or integrity.

## 11. Acknowledgement

We would like to express our gratitude to the Malaysian Ministry of Science, Technology, and Innovation (MOSTI) for funding this study.

## References

1. https://www.who.int/news-room/fact-sheets/detail/the-top-10-causes-of-death. The top 10 causes of death.

2. Vinicius de Araújo R, Silva Santos S, Missfeldt Sanches L, Giarolla J, El Seoud O, Igne Ferreira E. Malaria, and tuberculosis as diseases of neglected populations: state of the art in chemotherapy and advances in the search for new drugs. Mem Inst Oswaldo Cruz. 2020.

3. Ashley EA, Pyae Phyo A, Woodrow CJ. Malaria. Vol. 391, The Lancet. Lancet Publishing Group; 2018. p. 1608–21.

4. WHO. World malaria report 2022 [Internet]. 2022. Available from: https://www.who.int/teams/global-malaria-programme

5. Rossati A, Bargiacchi O, Kroumova V, Zaramella M, Caputo A, Garavelli PL. Climate, environment, and transmission of malaria. Infezioni in Medicina. 2016;24(2):93–104.

6. CDC. CDC - Malaria - Malaria Worldwide - Impact of Malaria. 2021.

7. WHO. Global Tuberculosis report 2022 [Internet]. 2022. Available from: http://apps.who.int/bookorders.

8. WHO. Artemisinin resistance and artemisinin-based combination therapy efficacy. 2018.

9. WHO. World malaria report 2016. 2016. 148 p.

10. Bell GJ, Goel V, Essone P, Dosoo D, Adu B, Mensah BA, et al. Malaria Transmission Intensity Likely Modifies RTS, S/AS01 Efficacy Due to a Rebound Effect in Ghana, Malawi, and Gabon. Journal of Infectious Diseases. 2022;226(9):1646–56.

11. Tameris M, Mearns H, Penn-Nicholson A, Gregg Y, Bilek N, Mabwe S, et al. Live-attenuated *Mycobacterium tuberculosis* vaccine MTBVAC versus BCG in adults and neonates: a randomised controlled, double-blind dose-escalation trial. Lancet Respir Med. 2019 Oct;7(9):757–70.

12. Ouédraogo A, Tiono AB, Kargougou D, Yaro JB, Ouédraogo E, Kaboré Y, et al. A phase 1b randomized, controlled, double-blinded dosage-escalation trial to evaluate the safety, reactogenicity and immunogenicity of an adenovirus type 35 based circumsporozoite malaria vaccine in burkinabe healthy adults 18 to 45 years of age. PLoS One. 2013 Oct;8(11).

13. Kimani D, Jagne YJ, Cox M, Kimani E, Bliss CM, Gitau E, et al. Translating the Immunogenicity of Prime-boost Immunization with ChAd63 and MVA ME-TRAP From Malaria Naive to Malaria-endemic Populations. Molecular Therapy. 2014;22(11):1992–2003.

14. Sirima SB, Durier C, Kara L, Houard S, Gansane A, Loulergue P, et al. Safety and immunogenicity of a recombinant *Plasmodium falciparum* AMA1-DiCo malaria vaccine adjuvanted with GLA-SE or Alhydrogel® in European and African adults: A phase 1a/1b, randomized, double-blind multi-centre trial. Vaccine. 2017 Oct;35(45):6218–27.

15. Dassah S, Adu B, Sirima SB, Mordmüller B, Ngoa UA, Atuguba F, et al. Extended follow-up of children in a phase2b trial of the GMZ2 malaria vaccine. Vaccine. 2021 Oct;39(31):4314–9.

16. Dejon-Agobe JC, Ateba-Ngoa U, Lalremruata A, Homoet A, Engelhorn J, Nouatin OP, et al. Controlled Human Malaria Infection of Healthy Adults with Lifelong Malaria Exposure to Assess Safety, Immunogenicity, and Efficacy of the Asexual Blood Stage Malaria Vaccine Candidate GMZ2. Clinical Infectious Diseases. 2019 Oct;69(8):1377–84.

17. Partnership. A Phase 3 Trial of RTS, S/AS01 Malaria Vaccine in African Infants. New England Journal of Medicine. 2012 Oct;367(24):2284–95.

18. Moncunill G, Rosa SC De, Ayestaran A, Nhabomba AJ, Mpina M, Cohen KW, et al. RTS, S/AS01E Malaria Vaccine Induces Memory and Polyfunctional T Cell Responses in a Pediatric African Phase III Trial. Front Immunol. 2017;8.

19. Otieno L, Oneko M, Otieno W, Abuodha J, Owino E, Odero C, et al. Safety and immunogenicity of RTS, S/AS01 malaria vaccine in infants and children with WHO stage 1 or 2 HIV disease: a randomised, double-blind, controlled trial. www.thelancet.com/infection [Internet]. 2016;16. Available from: 10.1016/

20. Rts SCTP, Tinto H, D’Alessandro U, Sorgho H, Valea I, Tahita MC, et al. Efficacy and safety of RTS, S/AS01 malaria vaccine with or without a booster dose in infants and children in Africa: final results of a phase 3, individually randomised, controlled trial. Lancet. 2015;386(9988):31–45.

21. Jongo SA, Shekalaghe SA, Church LWP, Ruben AJ, Schindler T, Zenklusen I, et al. Safety, immunogenicity, and protective efficacy against controlled human malaria infection of *Plasmodium falciparum* sporozoite vaccine in Tanzanian adults. American Journal of Tropical Medicine and Hygiene. 2018;99(2):338–49.

22. Sissoko MS, Healy SA, Katile A, Omaswa F, Zaidi I, Gabriel EE, et al. Safety and efficacy of PfSPZ Vaccine against *Plasmodium falciparum* via direct venous inoculation in healthy malaria-exposed adults in Mali: a randomised, double-blind phase 1 trial. Lancet Infect Dis. 2017 Oct;17(5):498–509.

23. Shekalaghe S, Rutaihwa M, Billingsley PF, Chemba M, Daubenberger CA, James ER, et al. Controlled human malaria infection of Tanzanians by intradermal injection of aseptic, purified, cryopreserved *Plasmodium falciparum* sporozoites. American Journal of Tropical Medicine and Hygiene. 2014 Oct;91(3):471–80.

24. Ndiaye BP, Thienemann F, Ota M, Landry BS, Camara M, Dièye S, et al. Safety, immunogenicity, and efficacy of the candidate tuberculosis vaccine MVA85A in healthy adults infected with HIV-1: A randomised, placebo-controlled, phase 2 trial. Lancet Respir Med. 2015 Oct;3(3):190–200.

25. Odutola AA, Owolabi OA, Owiafe PK, McShane H, Ota MOC. A new TB vaccine, MVA85A, induces durable antigen-specific responses 14 months after vaccination in African infants. Vaccine. 2012 Oct;30(38):5591–4.

26. Nemes E, Hesseling AC, Tameris M, Mauff K, Downing K, Mulenga H, et al. Safety and immunogenicity of newborn MVA85A vaccination and selective, delayed bacille calmette-guerin for infants of human immunodeficiency virus-infected mothers: A phase 2 randomized, controlled trial. Clinical Infectious Diseases. 2018 Feb 15;66(4):554–63.

27. Tameris MD, Hatherill M, Landry BS, Scriba TJ, Snowden MA, Lockhart S, et al. Safety and efficacy of MVA85A, a new tuberculosis vaccine, in infants previously vaccinated with BCG: a randomised, placebo-controlled phase 2b trial. Lancet. 2013;381(9871):1021–8.

28. Churchyard GJ, Snowden MA, Hokey D, Dheenadhayalan V, McClain JB, Douoguih M, et al. The safety and immunogenicity of an adenovirus type 35-vectored TB vaccine in HIV-infected, BCG-vaccinated adults with CD4+ T cell counts >350 cells/mm3. Vaccine. 2015 Oct;33(15):1890–6.

29. Kagina BMN, Tameris MD, Geldenhuys H, Hatherill M, Abel B, Hussey GD, et al. The novel tuberculosis vaccine, AERAS-402, is safe in healthy infants previously vaccinated with BCG, and induces dose-dependent CD4 and CD8T cell responses. Vaccine. 2014 Oct;32(45):5908–17.

30. Walsh DS, Owira V, Polhemus M, Otieno L, Andagalu B, Ogutu B, et al. Adenovirus type 35-vectored tuberculosis vaccine has an acceptable safety and tolerability profile in healthy, BCG-vaccinated, QuantiFERON®-TB Gold (+) Kenyan adults without evidence of tuberculosis. Vaccine. 2016 Oct;34(21):2430–6.

31. Bekker LG, Dintwe O, Fiore-Gartland A, Middelkoop K, Hutter J, Williams A, et al. A phase lb randomized study of the safety and immunological responses to vaccination with H4:IC31, H56:IC31, and BCG revaccination in *Mycobacterium tuberculosis*-uninfected adolescents in Cape Town, South Africa. EClinicalMedicine. 2020;21.

32. Idoko OT, Owolabi OA, Owiafe PK, Moris P, Odutola A, Bollaerts A, et al. Safety and immunogenicity of the M72/AS01 candidate tuberculosis vaccine when given as a booster to BCG in Gambian infants: An open label randomized controlled trial. Tuberculosis. 2014 Oct;94(6):564–78.

33. Meeren O Van Der, Hatherill M, Nduba V, Wilkinson RJ, Muyoyeta M, Brakel E Van, et al. Phase 2b Controlled Trial of M72/AS01 E Vaccine to Prevent Tuberculosis. New England Journal of Medicine. 2018 Oct;379(17):1621–34.

34. Penn-Nicholson A, Geldenhuys H, Burny W, van der Most R, Day CL, Jongert E, et al. Safety and immunogenicity of candidate vaccine M72/AS01E in adolescents in a TB endemic setting. Vaccine. 2015 Oct;33(32):4025–34.

35. Nell AS, D’Lom E, Bouic P, Sabaté M, Bosser R, Picas J, et al. Safety, tolerability, and immunogenicity of the novel antituberculous vaccine RUTI: Randomized, placebo-controlled phase II clinical trial in patients with latent tuberculosis infection. PLoS One. 2014 Oct;9(2).

36. Hesseling AC, Jaspan HB, Black GF, Nene N, Walzl G. Immunogenicity of BCG in HIV-exposed and non-exposed infants following routine birth or delayed vaccination. International Journal of Tuberculosis and Lung Disease. 2015 Oct;19(4):454–62.

37. Loxton AG, Knaul JK, Grode L, Gutschmidt A, Meller C, Eisele B, et al. Safety and immunogenicity of the recombinant mycobacterium bovis BCG vaccine VPM1002 in HIV-unexposed newborn infants in South Africa. Clinical and Vaccine Immunology. 2017 Oct;24(2).

38. Page MJ, Mckenzie JE, Bossuyt PM, Boutron I, Hoffmann TC, Mulrow CD, et al. Updating guidance for reporting systematic reviews: development of the PRISMA 2020 statement What is new? J Clin Epidemiol [Internet]. 2021; 134:103–12. Available from: 10.1016/j.jclinepi.2021.02.003

39. Tait DR, Hatherill M, Meeren O Van Der, Ginsberg AM, Brakel E Van, Salaun B, et al. Final Analysis of a Trial of M72/AS01 E Vaccine to Prevent Tuberculosis. New England Journal of Medicine. 2019 Oct;381(25):2429–39.

40. Tchakoute CT, Hesseling AC, Kidzeru EB, Gamieldien H, Passmore JAS, Jones CE, et al. Delaying BCG vaccination until 8 weeks of age results in robust BCG-specific T-cell responses in HIV-exposed infants. Journal of Infectious Diseases. 2015 Oct;211(3):338–46.

41. Tameris M, Hokey DA, Nduba V, Sacarlal J, Laher F, Kiringa G, et al. A double-blind, randomised, placebo-controlled, dose-finding trial of the novel tuberculosis vaccine AERAS-402, an adenovirus-vectored fusion protein, in healthy, BCG-vaccinated infants. Vaccine. 2015 Oct;33(25):2944–54.

42. Penn-Nicholson A, Tameris M, Smit E, Day TA, Musvosvi M, Jayashankar L, et al. Safety and immunogenicity of the novel tuberculosis vaccine ID93 + GLA-SE in BCG-vaccinated healthy adults in South Africa: a randomised, double-blind, placebo-controlled phase 1 trial. Lancet Respir Med. 2018 Oct;6(4):287–98.

43. Lutwama F, Kagina BM, Wajja A, Waiswa F, Mansoor N, Kirimunda S, et al. Distinct T-Cell Responses When BCG Vaccination Is Delayed from Birth to 6 Weeks of Age in Ugandan Infants. Journal of Infectious Diseases. 2014;209(6):887–97.

44. Nemes E, Geldenhuys H, Rozot V, Rutkowski KT, Ratangee F, Bilek N, et al. Prevention of M. tuberculosis Infection with H4:IC31 Vaccine or BCG Revaccination. New England Journal of Medicine. 2018 Oct;379(2):138–49.

45. Suliman S, Geldenhuys H, Johnson JL, Hughes JE, Smit E, Murphy M, et al. Bacillus Calmette– Guérin (BCG) Revaccination of Adults with Latent *Mycobacterium tuberculosis* Infection Induces Long-Lived BCG-Reactive NK Cell Responses. The Journal of Immunology. 2016 Oct;197(4):1100– 10.

46. Tameris MD, Hatherill M, Landry BS, Scriba TJ, Snowden MA, Lockhart S, et al. Safety and efficacy of MVA85A, a new tuberculosis vaccine, in infants previously vaccinated with BCG: A randomised, placebo-controlled phase 2b trial. The Lancet. 2013;381(9871):1021–8.

47. Geldenhuys HD, Mearns H, Foster J, Saxon E, Kagina B, Saganic L, et al. A randomized clinical trial in adults and newborns in South Africa to compare the safety and immunogenicity of bacille Calmette-Guérin (BCG) vaccine administration via a disposable-syringe jet injector to conventional technique with needle and syringe. Vaccine. 2015 Oct;33(37):4719–26.

48. Hatherill M, Geldenhuys H, Pienaar B, Suliman S, Chheng P, Debanne SM, et al. Safety and reactogenicity of BCG revaccination with isoniazid pretreatment in TST positive adults. Vaccine. 2014 Oct;32(31):3982–8.

49. Agnandji ST, Lell B, Fernandes JF, Abossolo BP, Kabwende AL, Adegnika AA, et al. Efficacy and Safety of the RTS, S/AS01 Malaria Vaccine during 18 Months after Vaccination: A Phase 3 Randomized, Controlled Trial in Children and Young Infants at 11 African Sites. PLoS Med. 2014;11(7).

50. Otieno L, Mendoza YG, Adjei S, Agbenyega T, Agnandji ST, Aide P, et al. Safety and immunogenicity of the RTS, S/AS01 malaria vaccine in infants and children identified as HIV-infected during a randomized trial in sub-Saharan Africa. Vaccine. 2020 Oct;38(4):897–906.

51. Bejon P, White MT, Olotu A, Bojang K, Lusingu JPA, Salim N, et al. Efficacy of RTS, S malaria vaccines: individual-participant pooled analysis of phase 2 data. Lancet Infectious Diseases. 2013;13(4):319–27.

52. Oneko M, Steinhardt LC, Yego R, Wiegand RE, Swanson PA, Natasha KC, et al. Safety, immunogenicity, and efficacy of PfSPZ Vaccine against malaria in infants in western Kenya: a double-blind, randomized, placebo-controlled phase 2 trial. Nat Med. 2021;27(9):1636-+.

53. Bell GJ, Loop MS, Mvalo T, Juliano JJ, Mofolo I, Kamthunzi P, et al. Environmental modifiers of RTS, S/AS01 malaria vaccine efficacy in Lilongwe, Malawi. BMC Public Health. 2020;20(1).

54. Dobaño C, Sanz H, Sorgho H, Dosoo D, Mpina M, Ubillos I, et al. Concentration and avidity of antibodies to different circumsporozoite epitopes correlate with RTS, S/AS01E malaria vaccine efficacy. Nat Commun. 2019 Oct;10(1).

55. Mendoza Y, Garric E, Leach A, Lievens M, Ofori-Anyinam O, Pirçon JY, et al. Safety profile of the RTS, S/AS01 malaria vaccine in infants and children: additional data from a phase III randomized controlled trial in sub-Saharan Africa. Hum Vaccin Immunother. 2019 Oct;15(10):2386–98.

56. Datoo MS, Natama MH, Some A, Traore O, Rouamba T, Bellamy D, et al. Efficacy of a low-dose candidate malaria vaccine, R21 in adjuvant Matrix-M, with seasonal administration to children in Burkina Faso: a randomised controlled trial. Lancet. 2021;397(10287):1809–18.

57. Berry AA, Gottlieb ER, Kouriba B, Diarra I, Thera MA, Dutta S, et al. Immunoglobulin G subclass and antibody avidity responses in Malian children immunized with *Plasmodium falciparum* apical membrane antigen 1 vaccine candidate FMP2.1/AS02(A). Malar J. 2019;18.

58. Han L, Hudgens MG, Emch ME, Juliano JJ, Keeler C, Martinson F, et al. RTS, S/AS01 Malaria Vaccine Efficacy is Not Modified by Seasonal Precipitation: Results from a Phase 3 Randomized Controlled Trial in Malawi. Sci Rep. 2017 Oct;7(1).

59. Mensah VA, Gueye A, Ndiaye M, Edwards NJ, Wright D, Anagnostou NA, et al. Safety, immunogenicity, and efficacy of prime-Boost vaccination with Chad63 and MVA encoding me-TRAP against *Plasmodium falciparum* infection in adults in Senegal. PLoS One. 2016 Oct;11(12).

60. Thera MA, Coulibaly D, Kone AK, Guindo AB, Traore K, Sall AH, et al. Phase 1 randomized controlled trial to evaluate the safety and immunogenicity of recombinant Pichia pastoris-expressed *Plasmodium falciparum* apical membrane antigen 1 (PfAMA1-FVO [25-545]) in healthy Malian adults in Bandiagara. Malar J. 2016 Oct;15(1).

61. Ubillos I, Ayestaran A, Nhabomba AJ, Dosoo D, Vidal M, Jiménez A, et al. Baseline exposure, antibody subclass, and hepatitis B response differentially affect malaria protective immunity following RTS, S/AS01E vaccination in African children. BMC Med. 2018 Oct;16(1).

62. Neafsey DE, Juraska M, Bedford T, Benkeser D, Valim C, Griggs A, et al. Genetic Diversity and Protective Efficacy of the RTS, S/AS01 Malaria Vaccine. New England Journal of Medicine. 2015 Oct;373(21):2025–37.

63. Gyaase S, Asante KP, Adeniji E, Boahen O, Cairns M, Owusu-Agyei S. Potential effect modification of RTS, S/AS01 malaria vaccine efficacy by household socio-economic status. BMC Public Health. 2021;21(1).

64. Chandramohan D, Zongo I, Sagara I, Cairns M, Yerbanga RS, Diarra M, et al. Seasonal Malaria Vaccination with or without Seasonal Malaria Chemoprevention. New England Journal of Medicine. 2021;385(11):1005–17.

65. Tinto H, Otieno W, Gesase S, Sorgho H, Otieno L, Liheluka E, et al. Long-term incidence of severe malaria following RTS, S/AS01 vaccination in children and infants in Africa: an open-label 3-year extension study of a phase 3 randomised controlled trial. Lancet Infect Dis. 2019 Aug 1;19(8):821–32.

66. Nouatin O, Ibáñez J, Fendel R, Ngoa UA, Lorenz FR, Dejon-Agobé JC, et al. Cellular and antibody response in GMZ2-vaccinated Gabonese volunteers in a controlled human malaria infection trial. Malar J. 2022 Dec 1;21(1).

67. Langowski MD, Khan FA, Savransky S, Brown DR, Balasubramaniyam A, Harrison WB, et al. Restricted valency (NPNA)n repeats and junctional epitope-based circumsporozoite protein vaccines against *Plasmodium falciparum*. NPJ Vaccines. 2022 Dec 1;7(1).

68. Suliman S, Luabeya AKK, Geldenhuys H, Tameris M, Hoff ST, Shi Z, et al. Dose optimization of H56:IC31 vaccine for tuberculosis-endemic populations a double-blind, placebo-controlled, dose-selection trial. Am J Respir Crit Care Med. 2019 Jan 15;199(2):220–31.

69. Geldenhuys H, Mearns H, Miles DJC, Tameris M, Hokey D, Shi Z, et al. The tuberculosis vaccine H4: IC31 is safe and induces a persistent polyfunctional CD4 T cell response in South African adults: A randomized controlled trial. Vaccine. 2015 Jul 9;33(30):3592–9.

70. Riste M, Marshall JL, Satti I, Harris SA, Wilkie M, Ramon RL, et al. Phase i trial evaluating the safety and immunogenicity of candidate tb vaccine mva85a, delivered by aerosol to healthy Mtb-infected adults. Vaccines (Basel). 2021;9(4).

71. Saveria T, Parthiban C, Seilie AM, Brady C, Martinez A, Manocha R, et al. Needle-free, spirulina-produced *Plasmodium falciparum* circumsporozoite vaccination provides sterile protection against pre-erythrocytic malaria in mice. NPJ Vaccines. 2022 Dec 1;7(1).

